# Signal Quality Screening and Automated Sleep Stage Agreement in Home EEG: A Systematic Comparison of Dreamento and YASA on the Wearanize+ Dataset

**DOI:** 10.64898/2026.06.01.26354591

**Authors:** Yildiz Dilara Parry, Giovanni Briganti

## Abstract

Wearable EEG devices such as the Zmax headband offer scalable alternatives to laboratory polysomnography (PSG) for sleep monitoring, but their real-world performance in home settings remains poorly characterised. This study presents a systematic validation of automated sleep staging on the Wearanize+ dataset; a unique multimodal resource providing synchronised full PSG, bilateral Zmax EEG (F7-Fpz/F8-Fpz), and psychiatric phenotyping from 100 participants recorded at home. We first developed and applied an automated signal quality screening framework, revealing that 10% of recordings failed completely due to signal dropout and a further 16% showed partial degradation. We then evaluated two automated staging algorithms; Dreamento and YASA against PSG manual scoring, stratified by signal quality. In technically adequate recordings (N=74), YASA achieved significantly higher agreement than Dreamento (mean κ=0.450 vs 0.371; Δκ=+0.079, p=0.0005), primarily through substantially improved N2 detection (recall: 0.64 vs 0.36). Both algorithms showed a systematic N2/N3 boundary confusion, however in opposite directions: Dreamento over-called N3 (37% of N2 epochs mis-staged as N3), while YASA over-called N2 (35% of N3 epochs mis-staged as N2). Critically, Dreamento showed greater robustness than YASA in degraded-quality recordings (WARN group: κ=0.414 vs 0.330), consistent with its training on Zmax-specific data. Signal quality metrics did not predict staging performance within adequate recordings, indicating that channel topology is the primary limiting factor for frontal single-channel staging. These findings establish the Wearanize+ dataset as a benchmark for wearable sleep staging and motivate the use of PSG manual stage labels for downstream physiological analyses.

## 1. Introduction

Sleep is a dynamically regulated biological state whose architecture, the temporal organisation of NREM and REM stages, slow-wave activity, and sleep spindle density, encodes clinically meaningful information about neural, metabolic, and psychiatric health. Polysomnography (PSG) remains the gold standard for sleep architecture assessment, providing simultaneous multi-channel EEG, EOG, EMG, ECG, and respiratory signals at clinical resolution. However, PSG is resource-intensive, ecologically constraining, and incompatible with population-scale longitudinal research: its throughput is limited to a small number of recordings per facility per night, and the clinical environment produces well-documented first-night effects that compromise ecological validity (Chinoy et al., 2021).

Consumer wearable EEG devices have emerged as a scalable alternative for at-home sleep monitoring. Among these, the Zmax headband (Hypnodyne Corp.) has attracted particular scientific interest due to its bilateral frontal EEG recording (F7-Fpz, F8-Fpz), open data format, and active research community. Unlike actigraphy-based wearables that infer sleep from movement, Zmax captures cortical EEG signals, theoretically enabling staging of NREM sub-stages and REM. However, this promise is contingent on two conditions that have not been systematically evaluated in large home-recording datasets: (1) adequate signal quality in unattended overnight home use, and (2) reliable automated staging from a non-standard frontotemporal referencing montage.

The Wearanize+ dataset provides the empirical conditions necessary to address both questions. Comprising synchronised full PSG, three concurrent wearable modalities, and psychiatric phenotyping from 130 healthy adults recorded in their own homes, it is the largest publicly available aligned PSG-Zmax dataset to date. The PlugNPlay version used in the present study includes 100 participants for whom both PSG and Zmax recordings were available with expert manual PSG scoring.

Two automated staging algorithms are currently used with Zmax data in the research community: DreamentoScorer (Dreamento), an open-source algorithm developed and validated specifically for Zmax; and YASA (Yet Another Spindle Algorithm), a widely used LightGBM-based staging algorithm trained on large-scale PSG cohorts. Despite their common use, no study has directly compared their performance on the Wearanize+ dataset with explicit signal quality stratification and systematic per-stage error analysis.

The present study addresses this gap with three specific aims: (1) develop and validate an automated signal quality screening framework applicable to unattended home EEG recordings, characterising recording failure rates and their consequences for staging performance; (2) provide the first direct head-to-head comparison of Dreamento and YASA on a large publicly available home-recording dataset with concurrent PSG ground truth and explicit signal quality stratification; and (3) characterise the systematic error patterns of each algorithm to identify the mechanistic basis of staging disagreement and derive practical recommendations for downstream physiological analyses. Beyond dataset-specific benchmarking, the study makes four contributions to the broader wearable EEG validation literature. First, it proposes and validates a signal quality stratification framework that is transferable to any home EEG study using frontal bipolar derivations, reframing the validation question from aggregate performance to quality-conditional performance; the operationally relevant metric for study design. Second, it identifies a quality-by-algorithm interaction not previously reported: YASA is superior in technically adequate recordings while Dreamento shows greater robustness in degraded signal, providing algorithm selection guidance that depends on anticipated recording quality rather than a single universal recommendation. Third, it characterises opposing N2/N3 boundary confusion patterns across algorithms, establishing the mechanistic basis for a key methodological recommendation: that downstream spectral and biomarker analyses should use PSG manual stage labels for epoch selection rather than automated wearable staging. Fourth, it establishes performance benchmarks on the largest publicly available aligned PSG-Zmax dataset to date, providing a reference against which future wearable EEG algorithms and datasets can be evaluated.

## 2. Methods

### 2.1 Dataset

We used the Wearanize+ (Sikder et al., 2026) PlugNPlay pre-processed dataset, comprising 100 participants (mean age 23.2 ± 4.3 years; 89 female) who underwent simultaneous at-home overnight recording with full PSG and the Zmax EEG headband. PSG was recorded using the SOMNOscreen Plus system (SOMNOmedics GmbH), with electrode placement and scoring according to AASM criteria (Iber, 2007). Manual PSG scoring was performed by a certified sleep scorer in 30-second epochs into five stages: Wake (W), N1, N2, N3, and REM. The Zmax headband recorded bilateral frontal EEG at 256 Hz from two bipolar derivations: F7-Fpz (left) and F8-Fpz (right). All recordings were performed in participants’ own homes without laboratory supervision. The dataset is available via the Radboud Data Repository under a signed Data Use Agreement (DUA) permitting non-commercial academic research use.

### 2.2 Signal Quality Screening

For each participant, the Zmax left EEG channel (F7-Fpz) was divided into consecutive 30-second epochs and the standard deviation (std) of each epoch was computed as an index of signal amplitude. Three quality categories were defined: epochs with std below 10 μV were classified as flat (signal dropout or electrode-skin contact failure)(Grosselin et al., 2019); epochs with std above 500 μV were classified as artefacted (movement artefact or electrode displacement)(Gao et al., 2022). Usable epochs were defined as those falling within the 10–500 μV range. Subjects were assigned quality flags based on the proportion of usable epochs: OK (≥75%), WARN (50–74%), or FAIL (<50%). The epoch at which sustained signal loss began (defined as the first epoch of a run of 10 or more consecutive flat epochs) was recorded as the signal loss onset epoch. A muscle-band power ratio threshold of 0.10 was applied as a secondary spectral quality indicator, consistent with the benchmark that spectral artefact contamination exceeding approximately 10% of total signal power meaningfully compromises EEG spectral feature reliability(Liu et al., 2018; Muthukumaraswamy, 2013). This threshold was additionally informed by the empirical distribution of muscle-band ratios in this dataset, where 0.10 corresponded to approximately 1.5 standard deviations above the median in OK-quality subjects (median=0.015, SD=0.058)

### 2.3 Automated Sleep Staging

**Dreamento:** DreamentoScorer is an open-source algorithm developed and validated specifically for the Zmax headband (Esfahani et al., 2023). Pre-computed Dreamento scores available within the Wearanize+ PlugNPlay dataset were used directly. Dreamento staging labels were encoded as integers 0-4 corresponding to Wake, N1, N2, N3, and REM respectively.

**YASA:** YASA (Vallat & Walker, 2021) version 0.6.5 was applied to all recordings. The Zmax F7-Fpz signal was converted from μV to Volts (multiplied by 10) and wrapped in an MNE RawArray object (MNE-Python version 1.8) with channel type EEG. YASA SleepStaging was initialised with the F7 channel as the primary EEG input and staging performed using the pre-trained LightGBM classifier. YASA output strings (W, N1, N2, N3, R) were mapped to the integer encoding scheme used for Dreamento and PSG.

YASA and Dreamento were selected as the primary algorithms because they represent the two principal open-source automated staging tools validated for frontal wearable EEG, with YASA trained on central PSG channels and Dreamento specifically designed for the Zmax F7-Fpz derivation. Evaluation of additional general-purpose classifiers was beyond the scope of this validation study, which aimed to characterise the performance of algorithms available to Zmax users rather than to benchmark the broader algorithm space.

### 2.4 Performance Metrics

Staging performance was evaluated at the subject level using three metrics: overall accuracy (proportion of correctly classified epochs), balanced accuracy (mean recall across all five stages, correcting for class imbalance), and Cohen’s kappa (κ, correcting for chance agreement). Kappa thresholds: <0.2 = slight, 0.21– 0.40 = fair, 0.41-0.60 = moderate, 0.61–0.80 = substantial, >0.80 = near-perfect (Landis & Koch, 1977). Performance was reported for the full sample (N=100) and stratified by quality group. Pooled per-stage precision, recall, and F1-score were computed across all OK subjects. Paired Wilcoxon signed-rank tests were used to compare per-subject kappa between algorithms. Spearman correlations between signal quality metrics and kappa were computed within the OK group. All analyses were conducted in Python 3.9 using scikit-learn, pandas, numpy, MNE-Python, YASA, and scipy.

## 3. Results

### 3.1 Signal Quality Distribution

Of the 100 Wearanize+ participants, 74 (74%) were classified as OK, 16 (16%) as WARN, and 10 (10%) as FAIL. The FAIL group showed two distinct subtypes: seven subjects had near-complete signal dropout from recording onset (median epoch std <1 μV, flat epoch percentage >67%), consistent with electrode contact failure; two showed mid-recording signal loss (signal loss onset epochs 28 and 415 respectively); and one showed persistent high-amplitude artefact (median std=525 μV), consistent with continuous motion artefact. The usable epoch distribution was bimodal, with failure cases clustered below 50% and successful recordings clustered above 80%, consistent with the binary nature of dry-electrode contact quality.

### 3.2 Staging Agreement: Dreamento

Across all 100 subjects, Dreamento achieved mean accuracy of 0.490 (±0.190), mean balanced accuracy of c0.499 (±0.155), and mean κ of 0.349 (±0.213). In OK subjects (N=74), mean κ improved to 0.371 (±0.189), with 43.2% of OK subjects achieving κ≥0.40 and 14.9% achieving κ≥0.60. The FAIL group showed near-chance agreement (κ=0.084±0.115). Within the OK group, kappa ranged from near-zero to 0.84, with signal quality metrics explaining none of this variance (usable epoch %: r=0.123, p=0.22; median EEG std: r=−0.193, p=0.055). The distribution of per subject κ across quality groups for both algorithms is shown un Figure 1 and Table 2 in detail.

**Figure 1.**
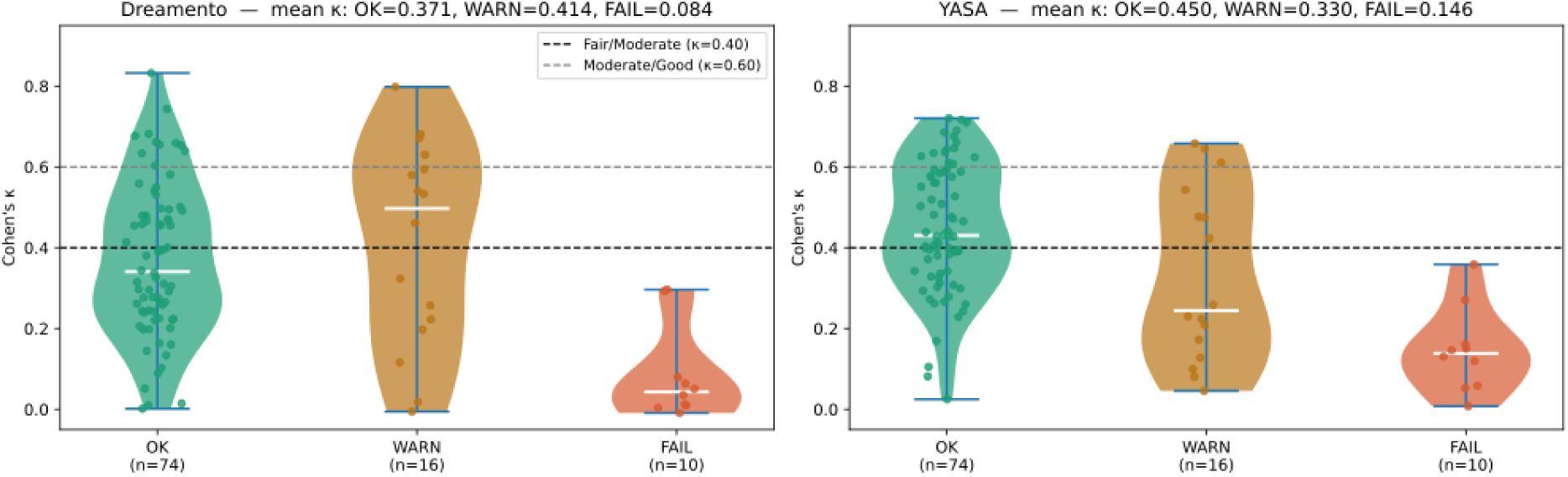
Distribution of Cohen’s κ for automated sleep staging: Dremento vs YASA (N=100)

**Table 1.** Signal quality distribution across the Wearanize+ PlugNPlay cohort (N=100).

| Quality group | N (%) | Usable epochs (%) | Flat epochs (%) | Median EEG std ($\mu\text{V}$) | Signal loss pattern |
| --- | --- | --- | --- | --- | --- |
| OK | 74 (74%) | $\geq 75\%$ | $<20\%$ | 20-50 | None or minimal |
| WARN | 16 (16%) | 50-74% | 15-47% | 10-90 | Partial, mid-recording |
| FAIL | 10 (10%) | $<50\%$ | $>67\%$ | $<1$ or $>500$ | Early or continuous |

**Table 2.** Staging performance stratified by signal quality: Dreamento vs YASA (N=100).

| Group | N | Dremento $\kappa$ mean<br>± SD [95% CI] | YASA $\kappa$ mean ±<br>SD [95% CI] | $\Delta\kappa$<br>(YASA–Dremento) |
| --- | --- | --- | --- | --- |
| All subjects | 100 | 0.349±0.213<br>[0.310–0.389] | 0.400±0.190<br>[0.365–0.438] | +0.051 (p=0.005 **) |
| OK only | 74 | 0.371±0.189<br>[0.328–0.413] | 0.450±0.160<br>[0.413–0.486] | +0.079 (p=0.0005 **) |
| OK + WARN | 90 | 0.379±0.201<br>[0.338–0.420] | 0.429±0.175<br>[0.394–0.464] | +0.050 (p=0.013 *) |
| FAIL only | 10 | 0.084±0.115<br>[0.026–0.157] | 0.146±0.104<br>[0.089–0.209] | +0.062 (p=0.13 ns) |
| WARN only | 16 | 0.414±0.253<br>[0.295–0.531] | 0.330±0.214<br>[0.234–0.430] | -0.084 (p=0.074 ns) |

95% CIs estimated by bootstrap resampling (2000 iterations).
\*\* p<0.01; \* p<0.05; ns = non-significant (Wilcoxon signed-rank test). WARN group comparison underpowered (post-hoc power analysis: N=22 required for 80% power at observed effect size d=0.601).

**Table 3.** Per-stage classification report: Dreamento vs YASA (OK subjects only, N=74).

| Stage | D prec | D rec | D F1 | Y prec | Y rec | Y F1 | Support |
| --- | --- | --- | --- | --- | --- | --- | --- |
| Wake | 0.27 | 0.88 | 0.41 | 0.31 | 0.85 | 0.46 | 7,164 |
| N1 | 0.25 | 0.12 | 0.16 | 0.14 | 0.11 | 0.13 | 3,956 |
| N2 | 0.89 | 0.36 | 0.51 | 0.75 | 0.64 | 0.69 | 33,018 |
| N3 | 0.45 | 0.80 | 0.58 | 0.90 | 0.50 | 0.64 | 13,356 |
| REM | 0.68 | 0.44 | 0.54 | 0.65 | 0.62 | 0.63 | 12,669 |
| Accuracy |  | 0.504 |  |  | 0.600 |  | 70,163 |

*D = Dreamento, Y = YASA. Prec = precision, rec = recall.*

### 3.3 Per-Stage Analysis: Dreamento

Per-stage analysis across OK subjects revealed systematic stage-depth overestimation. Wake was heavily over-predicted (recall=0.88, precision=0.27), absorbing 64% of true N1 and 43% of true REM epochs. N2 recall was severely impaired (0.36) with 37% of N2 epochs mis-staged as N3 (n=12,193), exceeding correct N2 classifications. N3 showed strong recall (0.80) but poor precision (0.45), reflecting the N2 to N3 over-calling. The normalised confusion matrices for both algorithms in OK-quality subjects are shown in Figure 2. The opposing N2/N3 boundary errors are clearly visible. REM recall was poor (0.44) with 43% called Wake. N1 detection was near-chance (recall=0.12). Epoch-level accuracy in OK subjects was 50.4%.

**Figure 2:**
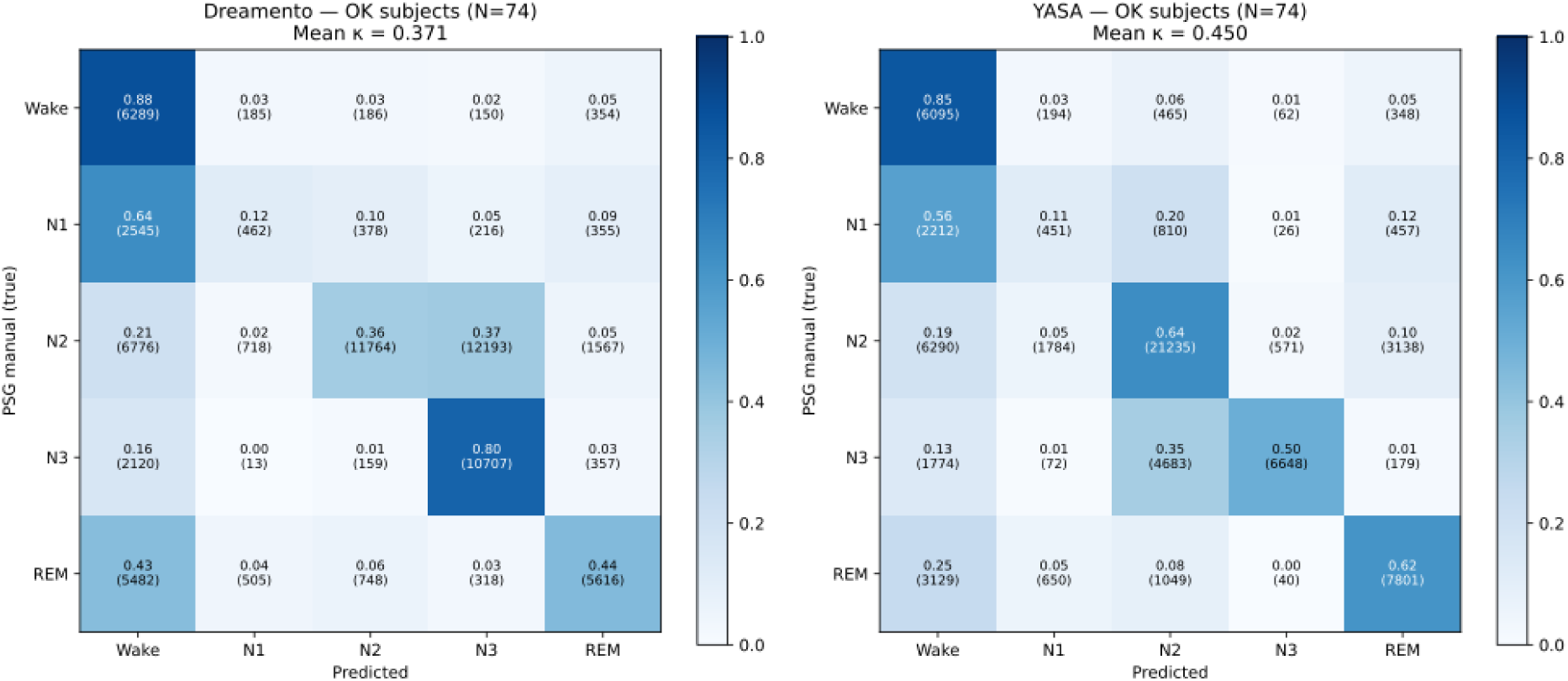
Normalised confusion matrices for Dreamento and YASA automated staging versus PSG manual scoring in OK-quality subjects (N=74).

### 3.4 Staging Agreement and Per-Stage Analysis: YASA

YASA achieved mean κ of 0.400 (±0.190) across all 100 subjects and 0.450 (±0.160) in OK subjects (N=74). YASA significantly outperformed Dreamento in OK subjects (paired Wilcoxon: Δκ=+0.079, p=0.0005) and across the full cohort (p=0.0051). Among OK subjects, 67 of 74 showed higher kappa with YASA than Dreamento. YASA achieved moderate agreement in the OK group (κ=0.450), crossing the fair-to-moderate boundary that Dreamento did not, as detailed on Figure 1.

On the other hand, the quality-by-algorithm interaction reversed in the WARN group: Dreamento outperformed YASA (mean κ=0.414 vs 0.330, Δκ=-0.084, p=0.074), consistent with Dreamento’s training on Zmax-specific recordings including degraded-quality data. This interaction was clearly visible in the per-subject scatter plot as Figure 3: green (OK) points clustered above the identity line (YASA better) while orange (WARN) points scattered at or below it.

**Figure 3.**
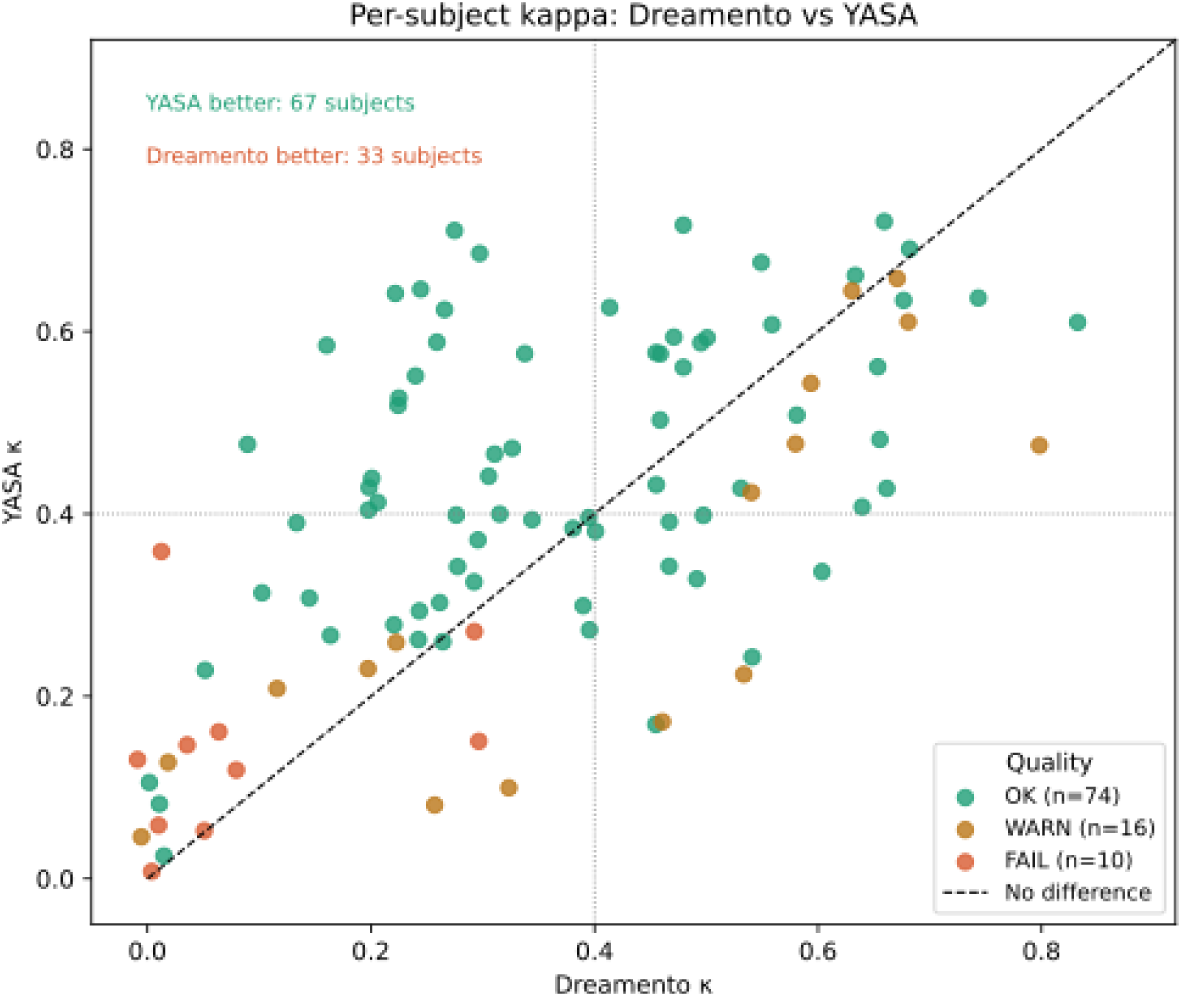
Per-subject Cohen’s κ for YASA versus Dreamento staging, stratified by signal quality group (N=100)

Per-stage analysis for YASA revealed a substantially different error pattern from Dreamento. N2 recall improved dramatically from 0.36 (Dreamento) to 0.64 (YASA), representing the primary source of overall kappa improvement. However, YASA introduced an opposing N2/N3 boundary error: 35% of true N3 epochs were classified as N2 (n=4,683), compared to Dreamento’s N2→N3 over-calling. YASA therefore trades Dreamento’s N3 over-calling for N2 over-calling, shifting rather than eliminating the N2/N3 boundary confusion. REM detection improved substantially (recall: 0.62 vs 0.44), attributable to YASA’s more conservative Wake threshold reducing the REM-Wake absorption. Wake over-prediction was reduced (recall 0.85 vs 0.88, precision improved from 0.27 to 0.31). N1 detection remained near-chance (recall=0.11), consistent across both algorithms and expected for single-channel frontal EEG. Epoch-level accuracy in OK subjects was 60.0%.

### 3.5 Left-Right Channel Comparison for YASA Staging

YASA automated staging was additionally evaluated on the right Zmax channel (F8-Fpz) to determine whether the bilateral design provides equivalent staging accuracy from either channel. The left channel showed significantly higher agreement with PSG manual scoring than the right channel (left κ =0.450±0.160 vs right κ =0.350±0.168; Wilcoxon signed-rank W=196, p<0.0001; rank-biserial r=0.859), with 88% of subjects (65/74) showing higher kappa on the left channel. The mean kappa advantage of the left channel was 0.100 (SD=0.113), representing a clinically meaningful improvement in staging agreement. This systematic left-channel advantage likely reflects preferential electrode-skin contact quality during home recordings, as right-side sleeping; the more prevalent sleep position in the general population positions the left Zmax electrode in closer contact with the forehead surface. These findings recommend the left channel (F7-Fpz) as the preferred input for automated staging algorithms when using the Zmax headband in home recording contexts.

## 4. Discussion

### 4.1 Signal quality in home EEG recording

The signal quality screening framework developed in this study revealed a 10% outright failure rate in the Wearanize+ home-recording cohort, consistent with published estimates for dry-electrode home EEG. Failure patterns were not random: seven of ten FAIL subjects showed near-zero amplitude from recording onset, suggesting inadequate initial electrode placement rather than contact loss during sleep. A brief signal quality check immediately before lights-off would likely prevent the majority of recording failures at negligible cost. The bimodal usable-epoch distribution, with few subjects in the 50–75% intermediate range, suggests that dry-electrode home EEG recording either succeeds or fails rather than degrading gradually, consistent with the binary nature of electrode-skin impedance.

A critical finding was that signal quality metrics did not significantly predict staging performance within the OK group (usable epoch %: r=0.123, p=0.22). This indicates that once minimum amplitude thresholds are met, staging disagreement is driven by channel topology rather than signal quality. The F7-Fpz and F8-Fpz bipolar derivations lack centroparietal coverage essential for AASM staging; sigma band spindle detection at Cz, occipital alpha for Wake, and chin EMG for REM confirmation. No degree of improved frontal signal quality can compensate for the absence of these channels, and algorithm development must therefore focus on improving spectral discrimination from frontal EEG rather than on signal quality optimisation alone.

### 4.2 YASA outperforms Dreamento in adequate-quality recordings

YASA achieved significantly higher staging agreement than Dreamento in technically adequate recordings (κ=0.450 vs 0.371, p=0.0005), with 67 of 74 OK subjects showing higher individual kappa with YASA. The primary source of improvement was N2 recall, which nearly doubled from 0.36 (Dreamento) to 0.64 (YASA). This improvement is attributable to YASA’s LightGBM classifier being trained on a large, diverse PSG dataset in which N2 is the dominant sleep stage where the model is therefore well-calibrated to the spectral characteristics of N2, including the sigma-band activity from sleep spindles and the slower delta components of K-complexes, even when observed through a frontal rather than central channel. REM detection also improved substantially (recall: 0.62 vs 0.44), reflecting YASA’s more conservative Wake threshold reducing the erroneous Wake absorption of REM epochs.

Both algorithms showed N2/N3 boundary confusion, but in opposing directions. Dreamento over-called N3 (37% of N2 classified as N3), while YASA over-called N2 (35% of N3 classified as N2). This opposing pattern is mechanistically consistent with their training origins: Dreamento, trained on Zmax forehead EEG where frontal slow-wave prominence makes the N2/N3 boundary particularly ambiguous, appears to use a liberal N3 threshold that overshoots; YASA, trained on standard PSG where N3 is defined by high delta power at central and parietal electrodes, applies a conservative N3 threshold relative to what is observable at frontal derivations. Neither algorithm resolves the fundamental ambiguity of the N2/N3 boundary from frontal EEG alone.

For applications requiring reliable epoch-level staging, such as stage-specific spectral biomarker extraction, the present results recommend YASA as the preferred algorithm for quality-adequate Zmax recordings with Dreamento reserved for degraded signal conditions where it demonstrates a compensatory advantage of approximately 45 epochs per night (22.6 minutes). The 9.8 percentage point accuracy advantage of YASA over Dreamento in quality-adequate recordings (60.2% vs 50.4%) corresponds to approximately 93 fewer misclassified epochs per night (46.4 minutes), which is clinically meaningful for stage-specific physiological analyses but attenuated for aggregate sleep architecture metrics where misclassification errors partially cancel across epochs.

For context, USleep pre-computed scores available in the Wearanize+ dataset; applied to the full PSG montage achieved mean κ=0.761 (±0.092) against manual scoring across OK-quality subjects (N=71), consistent with published multi-channel PSG staging benchmarks (Perslev et al., 2021) and quantifying the performance gap attributable to the absence of central, occipital, and EMG channels in single-channel frontal wearable recordings. The 0.311 kappa difference between USleep on PSG (κ=0.761) and YASA on Zmax (κ=0.450) represents the topological ceiling imposed by frontal-only EEG, independent of algorithm quality.

### 4.3 Dreamento is more robust to degraded signal quality

The quality-by-algorithm interaction is one of the most practically important findings of this study. Dreamento outperformed YASA in WARN subjects (mean κ=0.414 vs 0.330), reversing the OK-group advantage. This pattern is consistent with Dreamento’s training on Zmax-specific real-world recordings, which would include examples of partial signal degradation typical of home recording. YASA’s LightGBM classifier, trained on clean PSG data, has not encountered the spectral signatures of degraded dry-electrode frontal EEG and therefore performs less reliably when signal quality departs from the clean recording assumption. This has a practical implication for Zmax research: YASA should be preferred when signal quality is confirmed adequate (OK group), while Dreamento may be more appropriate for datasets with higher proportions of degraded recordings or when signal quality screening is not feasible. In the absence of signal quality information, the two algorithms perform comparably overall (κ=0.349 vs 0.400 across all subjects).

### 4.4 Implications for downstream physiological analyses

The N2/N3 boundary confusion in both algorithms has a specific and consequential implication for downstream physiological analyses. If automated staging labels are used to select epochs for feature extraction. For example, extracting sleep spindles from N2 epochs or computing slow oscillation amplitude from N3 epochs, the systematic misclassification will contaminate feature distributions. Dreamento’s N2-N3 over-calling means that Dreamento-labelled N3 epochs will contain substantial N2 sigma-band activity, inflating N3 spindle counts and reducing N3 delta dominance. YASA’s reverse pattern will correspondingly inflate N2 delta features. Both effects would confound any analysis that depends on stage-specific physiological features.

These findings strongly motivate the use of PSG manual stage labels as the epoch-selection ground truth for any downstream physiological analysis of Zmax data, reserving automated staging for studies where concurrent PSG is unavailable. In the Wearanize+ dataset, concurrent PSG is available for all 100 subjects, making PSG-referenced feature extraction feasible and methodologically appropriate. This design choice where Zmax provides the EEG signal, PSG provides the staging ground truth, is adopted in the planned follow-on spectral validation and psychiatric biomarker studies described below.

### 4.5 Future directions

Three research directions follow directly from the present findings. First, spectral validation of Zmax EEG against concurrent PSG within PSG-defined stage epochs, comparing band power correspondence (delta, sigma, alpha, beta), sleep spindle detection calibration between Zmax frontal and PSG central channels, and slow oscillation amplitude agreement. The concurrent PSG design of Wearanize+ converts the limitations identified here into calibration opportunities: systematic per-subject offsets between Zmax frontal and PSG central sigma power, for example, can be estimated and corrected rather than treated as fundamental barriers.

Second, development of multi-omics integration frameworks; specifically multi-omics factor analysis (MOFA+) and other multi-modal networks in order to learn personalised sleep biological representations from aligned PSG and wearable data. The staging validation reported here establishes a key methodological prerequisite: PSG manual stage labels should be used as epoch-selection ground truth for feature extraction in multi-omics analyses, ensuring that the biological representations learned reflect genuine sleep physiology rather than algorithmic staging artefacts.

### 4.6 Limitations

Several limitations should be noted. First, the Wearanize+ cohort comprises healthy young adults (18–39 years), limiting generalisability to clinical populations and older adults. Second, the single-night design precludes assessment of night-to-night variability in signal quality and staging agreement. Third, the signal quality screening framework uses amplitude-based criteria that may not detect frequency-specific artefacts preserving amplitude but distorting spectral content.Fourth, post-hoc spectral quality analysis revealed that 16.2% of amplitude-adequate recordings (12/74 OK subjects) showed muscle-band power ratios exceeding 10% of total power, indicating frequency-specific artefact contamination not captured by amplitude screening. Future studies using the Wearanize+ dataset should consider implementing a secondary spectral quality filter; for example, excluding subjects with muscle-band ratios above a defined threshold, to identify recordings where YASA’s spectral features may be unreliable despite adequate overall signal amplitude. Fifth, algorithms specifically trained on forehead EEG (ezscore-f) were not evaluated due to installation incompatibilities; future work should include this comparison.

## 5. Conclusions

This study provides the first systematic signal quality and staging validation of the Zmax EEG headband on the Wearanize+ home-recording dataset across 100 participants with concurrent PSG ground truth, and makes four contributions to the broader wearable EEG validation literature. Ten percent of home recordings failed due to signal dropout detectable by automated amplitude screening; a failure rate that has direct implications for sample size planning in home EEG studies. In technically adequate recordings, YASA significantly outperformed Dreamento (mean κ=0.450 vs 0.371, p=0.0005), primarily through improved N2 detection, while Dreamento showed greater robustness in degraded-quality recordings. This quality-by-algorithm interaction, not previously reported in the Zmax validation literature means that algorithm selection should be conditioned on anticipated recording quality rather than on aggregate performance benchmarks. Both algorithms showed opposing N2/N3 boundary confusion patterns, motivating the recommendation to use PSG manual stage labels for epoch selection in downstream spectral and biomarker analyses; a finding that directly shaped the design of the companion spectral validation and psychiatric biomarker studies in this programme. The signal quality screening framework, algorithm comparison methodology, and systematic error characterisation approach developed here are transferable to other home EEG datasets and wearable platforms using frontal bipolar derivations, providing a replicable template for wearable EEG validation studies. These findings establish the validated methodological prerequisites for the multi-paper programme of wearable-based sleep biomarker research of which this study is the first component.

## Data Availability Statement

The Wearanize+ PlugNPlay dataset used in this study is publicly available via the Radboud Data Repository under a signed Data Use Agreement permitting non-commercial academic research use. The dataset can be accessed at https://doi.org/10.1093/sleepadvances/zpaf094 (Sikder et al., 2025). Analysis code used to produce the results reported in this manuscript is available on request from the corresponding author.

## IRB Statement

This study used a publicly available secondary dataset (Wearanize+ PlugNPlay, Radboud Data Repository). All data collection, participant recruitment, and ethical oversight were conducted by the original dataset creators. The original study was approved by the relevant institutional ethics committee and all participants provided written informed consent prior to data collection, as described in Sikder et al. (2025). Secondary analysis of this anonymised publicly available dataset under a signed Data Use Agreement did not require additional ethical approval from the authors’ institutional review board.

## References

Chinoy, E. D., Cuellar, J. A., Huwa, K. E., Jameson, J. T., Watson, C. H., Bessman, S. C., Hirsch, D. A., Cooper, A. D., Drummond, S. P. A., & Markwald, R. R. (2021). Performance of seven consumer sleep-tracking devices compared with polysomnography. Sleep, 44(5), zsaa291. 10.1093/sleep/zsaa291

Esfahani, M. J., Weber, F. D., Boon, M., Anthes, S., Almazova, T., Hal, M. V., Keuren, Y., Heuvelmans, C., Simo, E., Bovy, L., Adelhöfer, N., Avest, M. M. T., Perslev, M., Horst, R. T., Harous, C., Sundelin, T., Axelsson, J., & Dresler, M. (2023). Validation of the sleep EEG headband ZMax. Neuroscience. 10.1101/2023.08.18.553744

Gao, Z., Cui, X., Wan, W., Qin, Z., & Gu, Z. (2022). Signal Quality Investigation of a New Wearable Frontal Lobe EEG Device. Sensors, 22(5), 1898. 10.3390/s22051898

Grosselin, F., Navarro-Sune, X., Vozzi, A., Pandremmenou, K., De Vico Fallani, F., Attal, Y., & Chavez, M. (2019). Quality Assessment of Single-Channel EEG for Wearable Devices. Sensors, 19(3), 601. 10.3390/s19030601

Iber, C. (2007). The AASM manual for the scoring of sleep and associated events: Rules, terminology, and technical specification. *(No Title)*.

Landis, J. R., & Koch, G. G. (1977). The Measurement of Observer Agreement for Categorical Data. Biometrics, 33(1), 159. 10.2307/2529310

Liu, J., Ramakrishnan, S., Laxminarayan, S., Neal, M., Cashmere, D. J., Germain, A., & Reifman, J. (2018). Effects of signal artefacts on electroencephalography spectral power during sleep: Quantifying the effectiveness of automated artefact rejection algorithms. Journal of Sleep Research, 27(1), 98–102. 10.1111/jsr.12576

Muthukumaraswamy, S. D. (2013). High-frequency brain activity and muscle artifacts in MEG/EEG: A review and recommendations. Frontiers in Human Neuroscience, 7. 10.3389/fnhum.2013.00138

Perslev, M., Darkner, S., Kempfner, L., Nikolic, M., Jennum, P. J., & Igel, C. (2021). U-Sleep: Resilient high-frequency sleep staging. Npj Digital Medicine, 4(1), 72. 10.1038/s41746-021-00440-5

Sikder, N., Verkaar, L., Paltarzhytskaya, A., Acan, S., Bovy, L., Almazova, T., Krugliakova, E., Rosenblum, Y., Krauledat, M., Dresler, M., & Zerr, P. (2026). *Wearanize+*: A multimodal dataset for evaluating wearable technologies in sleep research. SLEEP Advances, 7(1), zpaf094. 10.1093/sleepadvances/zpaf094

Vallat, R., & Walker, M. P. (2021). An open-source, high-performance tool for automated sleep staging. eLife, 10, e70092. 10.7554/eLife.70092

